# Outcome of SARS-CoV-2 infection linked to MAIT cell activation and cytotoxicity: evidence for an IL-18 dependent mechanism

**DOI:** 10.1101/2020.08.31.20185082

**Authors:** H. Flament, M. Rouland, L. Beaudoin, A. Toubal, L. Bertrand, S. Lebourgeois, Z. Gouda, C. Rousseau, P. Soulard, M. Hurtado-Nedelec, S. Luce, K. Bailly, M. Andrieu, C. Boitard, A. Vallet-Pichard, JF. Gautier, N. Ajzenberg, B. Terrier, F. Pene, J. Ghosn, Y. Yazdanpanah, B. Visseaux, D. Descamps, JF. Timsit, R.C. Monteiro, A. Lehuen

## Abstract

Immune system dysfunction is paramount in Coronavirus disease 2019 (COVID-19) severity and fatality rate. Mucosal-Associated Invariant T (MAIT) cells are innate-like T cells involved in mucosal immunity and protection against viral infections. Here, we studied the immune cell landscape, with emphasis on MAIT cells, in a cohort of 182 patients including patients at various stages of disease activity. A profound decrease of MAIT cell counts in blood of critically ill patients was observed. These cells showed a strongly activated and cytotoxic phenotype that positively correlated with circulating pro-inflammatory cytokines, notably IL-18. MAIT cell alterations markedly correlated with disease severity and patient mortality. SARS-CoV-2-infected macrophages activated MAIT cells in a cytokine-dependent manner involving an IFNα-dependent early phase and an IL-18-induced later phase. Therefore, altered MAIT cell phenotypes represent valuable biomarkers of disease severity and their therapeutic manipulation might prevent the inflammatory phase involved in COVID-19 aggravation.

## Introduction

The Severe Acute Respiratory Syndrome Coronavirus 2 (SARS-CoV-2) is the etiologic agent responsible for the recent outbreak of Coronavirus disease 2019 (Covid-19) that started in December 2019. Cellular targets of the SARS-CoV-2 are primarily upper- and lower respiratory tract cells as well as pulmonary cells^1,2^. The SARS-CoV-2 infection results in a wide range of clinical signs, from asymptomatic to life-threatening Acute Respiratory Distress Syndrome (ARDS), caused by a deleterious anti-viral immune response in the lung^3-5^. In serious cases, the overbalanced local immune response damages the airways and may lead to noncardiogenic pulmonary edema, hypoxia, and need for artificial ventilation and/or oxygenation^6,7^.

Innate-like T cells including Mucosal-associated invariant T (MAIT), invariant Natural Killer T (iNKT), and γδT cells, are known to be key actors of pulmonary mucosal immunity, of mucosal tissue repair and are involved in the immune response to numerous respiratory pathogens^8-13^. Innate Lymphoid cells (ILCs) have also emerged as important mediators in tissue protection and repair during lung viral infection^14,15^. Among them, MAIT cells recognize bacterial metabolites derived from the riboflavin synthesis pathway and presented by the major histocompatibility complex (MHC) class-I-related protein (MR1)^16,17^. It has been established that MAIT cells are activated during viral infections, especially in blood and lungs^18-20^. MAIT cell activation by viruses is TCR-independent and cytokine-dependent^18,20^. During both acute and chronic viral infections, MAIT cell blood frequency is reduced while expression of HLA-DR, PD-1, CD38 and CD69 is upregulated^18-20,22-24^. Upon acute viral infection, they produce high levels of granzyme B (GzB)^20^.

In COVID-19 patients, alteration of peripheral lymphocyte and myeloid subsets is associated with clinical characteristics and treatment efficiency^25-27^. However, status of MAIT cells, remain unknown in COVID-19 patients. Here, we analyzed blood MAIT cells of COVID-19 patients with different disease severity status from Infectious Disease Unit (IDU) (n=51) and Intensive Care Unit (ICU) (n=51). These patients were compared with uninfected controls (n=80) matched for age, sex and comorbidities. In COVID-19 patients, blood MAIT cells are altered in relation with disease severity and mortality. Macrophages infected *in vitro* with SARS-CoV-2 stimulate MAIT cells. Our study supports MAIT cells as a new indicator of COVID-19 gravity and suggest them as target to reduce COVID-19 morbidity.

## Results

### Adaptive and innate T cell frequency in the blood of COVID-19 patients

We first began our study of immune cells in SARS-CoV-2 infection by analyzing the frequency and phenotype of lymphocytes in blood samples from COVID-19 patients as well as (age-matched BMI-matched) non-infected donors (**Fig. 1a**). Fifty-one COVID-19 patients had been admitted in IDU (moderate cases) and 51 patients in ICU (severe cases) (with a death rate of 41% in ICU). As controls, we included 80 healthy non-infected donors as well as donors with various pathologies (diabetes, obesity) to match those affecting hospitalized COVID-19 patients (**Fig. 1a**). All characteristics and health data of recruited patients and controls are listed in **Table 1**. Whole blood was collected and stained for flow cytometry to analyze the frequencies of both innate and adaptive T lymphocytes following the gating strategy explained in **Supplementary Figure 1**. A majority of patients with COVID-19 presented a significant reduction of T cells in IDU, more pronounced in ICU patients (**Fig. 1b**), confirming earlier reports^28,29^. Among CD3^+^ T cells, there was a slight increase of blood αβ T cells frequency in ICU patients, mirrored by a decrease of γδT cells (**Fig. 1c**). Both CD4^+^ and CD8^+^ T cell frequency among CD3^+^ T cells were not significantly impacted by SARS-CoV-2 infection whereas T regulatory cell frequency was significantly reduced in both IDU and ICU patients compared to uninfected controls (**Fig. 1d**). We observed a collapse of Vα7.2^+^ CD161^+^ MAIT cell frequency among CD3^+^ cells in COVID-19 patients, down to a tenth of those observed in uninfected controls (**Fig. 1e**). We confirmed that MAIT cell markers CD161 and Vα7.2 allowed similar MAIT cell identification as with MR1 tetramers loaded with the active ligand 5-(2-oxopropylideneamino)-6-D-ribitylaminouracil (5-OP-RU) in all three groups of patients (**Supplementary Fig. 2**). Among MAIT cells, the CD8^+^ subset was further reduced in COVID-19 patients, but to a lesser extent in ICU compared to IDU (**Fig. 1f**). We therefore observed a dramatic decrease of blood MAIT cells in COVID-19 patients, with an alteration of MAIT cell subsets repartition.

**Table 1:** COVID-19 and uninfected patients clinical characteristics.

| Group | Uninfected controls | IDU | ICU |
| --- | --- | --- | --- |
| Number of patients | 80 | 51 | 51 |
| Sex |  |  |  |
| Female | 40% (32) | 33% (17) | 18% (9) |
| Male | 60% (48) | 67% (34) | 82% (42) |
| Age |  |  |  |
| Median (MAD) | 58 ±20.9 | 61 ±14.8 | 59 ±14.8 |
| Mean (SD) | 54.9 ±17.5 | 61 ±16.4 | 57.6 ±13.3 |
| Range | 23-78.8 | 23-98 | 29-83 |
| BMI |  |  |  |
| Median (MAD) | 28 ±4.6 | 30 ±5.3 | 29 ±5.9 |
| Mean (SD) | 27.2 ±4.9 | 28.9 ±5.3 | 29.8 ±5.4 |
| Range | 18-38.3 | 17-38 | 21-43 |
| Hospital |  |  |  |
| Bichat | 32% (26) | 86% (44) | 94% (48) |
| Cochin | 26% (21) | 14% (7) | 5.9% (3) |
| Beaujon-Lariboisière | 26% (21) | 0% (0) | 0% (0) |
| EFS | 15% (12) | 0% (0) | 0% (0) |
| Disease duration (days) |  |  |  |
| Median (MAD) | - | 10 ±5.9 | 18.5 ±13.3 |
| Mean (SD) | - | 13.5 ±11.9 | 24.5 ±16.7 |
| Range | - | 0-67 | 4-65 |
| Deaths | 0% (0) | 12% (6) | 41% (21) |
| Treatments |  |  |  |
| Tocilizumab | 0% (0) | 2% (1) | 12% (6) |
| Anakinra | 0% (0) | 12% (6) | 33% (17) |
| Lopinavir/Ritonavir | 0% (0) | 33% (17) | 57% (29) |
| Anti-IFNβ | 0% (0) | 2% (1) | 3.9% (2) |
| Dexamethasone | 0% (0) | 20% (10) | 67% (34) |
| Hydroxychloroquine | 0% (0) | 5.9% (3) | 18% (9) |
| Pulmonary lesions |  |  |  |
| Mild (score = 1) | 0% (0) | 12% (6) | 3.9% (2) |
| Moderate (score = 2) | 0% (0) | 33% (17) | 5.9% (3) |
| Severe (score = 3) | 0% (0) | 27% (14) | 24% (12) |
| Grievous (score = 4) | 0% (0) | 12% (6) | 20% (10) |
| Critical (score = 5) | 0% (0) | 0% (0) | 3.9% (2) |
| Missing | 100% (80) | 16% (8) | 43% (22) |
| Pulmonary lesions score |  |  |  |
| Median (MAD) | - | 2 ±1.5 | 3 ±1.5 |
| Mean (SD) | - | 2.5 ±0.9 | 3.2 ±1 |
| Range | - | 1-4 | 1-5 |
| PaO<sub>2</sub>/FiO<sub>2</sub> |  |  |  |
| Median (MAD) | - | 306.5 ±160.1 | 158 ±108.2 |
| Mean (SD) | - | 345.3 ±176.9 | 194.2 ±144.8 |
| Range | - | 88-700 | 55-948 |
| SAPS II score |  |  |  |
| Median (MAD) | - | 22 ±0 | 35 ±16.3 |
| Mean (SD) | - | 28.4 ±17.9 | 37.3 ±15.9 |
| Range | - | 16-60 | 9-78 |
| Polymorphonuclear neutrophils (PMN) |  |  |  |
| Median (MAD) | 4.2 ±1.9 | 3.7 ±2.2 | 9.7 ±4.8 |
| Mean (SD) | 4.3 ±1.8 | 4.4 ±2.7 | 10.6 ±6 |
| Range | 2.1-8.5 | 1.1-15.7 | 2.9-30.2 |
| C-Reactive protein (CRP) |  |  |  |
| Median (MAD) | 1.5 ±1.2 | 66 ±60.8 | 95.2 ±93.4 |
| Mean (SD) | 2.6 ±2.1 | 72.2 ±59.3 | 125.7 ±97.6 |
| Range | 0.2-6.9 | 3-241 | 6-428 |
| Procalcitonin (PCT) |  |  |  |
| Median (MAD) | - | 0.1 ±0.1 | 0.3 ±0.3 |
| Mean (SD) | - | 3.3 ±12.9 | 1.5 ±4.1 |
| Range | - | 0-55 | 0-18 |
| Type 2 Diabetes | 38% (30) | 27% (14) | 18% (9) |
| HbA1c |  |  |  |
| Median (MAD) | 7.4 ±0.9 | 7.7 ±1.9 | 6.6 ±0.7 |
| Mean (SD) | 7.6 ±1 | 8.1 ±2.4 | 6.8 ±1.2 |
| Range | 5.3-10.2 | 5.1-16.1 | 5.1-10.4 |
| Secondary Infection | 0% (0) | 5.9% (3) | 51% (26) |
| Hospital acquired pneumonia | 0% (0) | 4% (2) | 29.4% (15) |
| Septic shock | 0% (0) | 2% (1) | 17.6% (9) |

**Figure 1.**
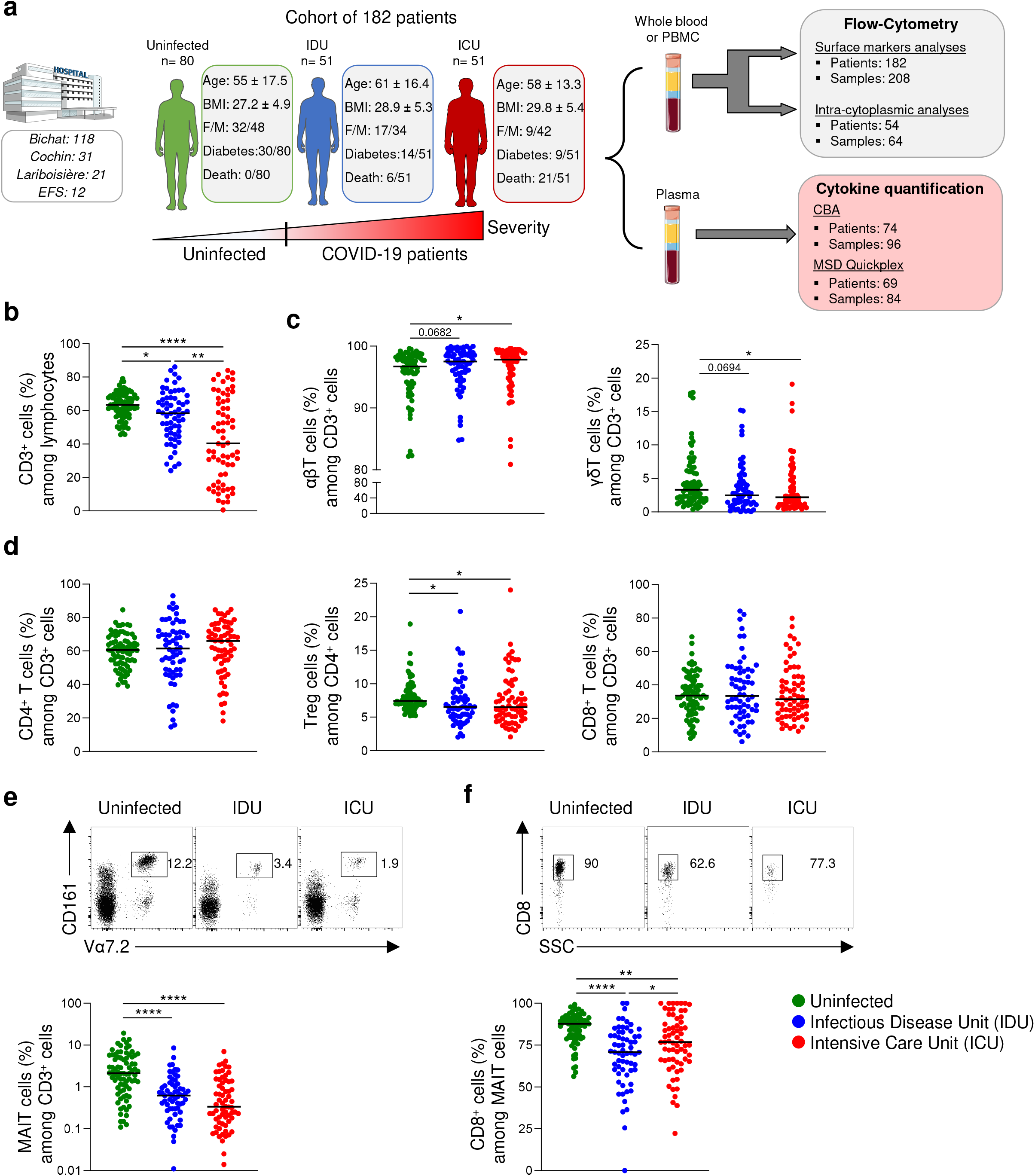
Immune cell frequencies and status in the blood of COVID-19 patients. (**a**) Graphical representation of the COVID-19 cohort including a total of 182 patients from different hospitals and described as following: age, BMI, sex (F= Female, M= Male), diabetes, and fatality rates. Mean (± SD) values for each medical ward are represented. (**b-c**) Whole blood or Peripheral Blood Mononuclear Cells (PBMC) were collected from patients and analyzed through flow cytometry. Flow cytometry analysis of CD3^+^ cells (**b**), αβT cells and γδT cells frequencies (**c**) from uninfected controls (n=80), COVID-19 patients hospitalized in an Infectious Disease Unit (IDU) (n=62) or hospitalized in an Intensive Care Unit (ICU) (n=66). (**d**) Flow cytometry analysis of CD4^+^ cells, regulatory T cells (Treg), and CD8^+^ T cell frequencies in the blood of patients as described in (**b-c**). (**e-f**) Representative dot plots of Vα7.2 and CD161 staining to identify MAIT cells in blood of uninfected control and COVID-19 patients from IDU and ICU, and MAIT cell (**e**) and CD8^+^ MAIT cell (**f**) frequencies in the blood of patients as described in (**b-c**). SSC, side scatter. Small horizontal lines indicate the median, each symbol represents one biological sample. *P<0.05, **P<0.01, P<0.001, and *P<0.0001 (two-sided Mann-Whitney nonparametric test).

### MAIT cells are activated and cytotoxic in COVID-19 patients

As MAIT cell frequency is extremely reduced in blood of COVID-19 patients, we analyzed both their surface markers expression as well as their cytokine production. An activated phenotype with a dramatic increase of CD69 expression was observed in COVID-19 patients, with a median of 41% CD69^+^ MAIT cells in IDU and 67% in ICU patients, with some reaching 100% (**Fig. 2a**). Blood MAIT cells also displayed a significant increase of the NK cell-associated activation CD56^+^ marker compared to controls, that was highest in ICU patients (**Fig. 2a**). Accordingly, double positive CD56^+^ CD69^+^ MAIT cell frequency was increased in IDU patients and even more in ICU patients (**Fig. 2a**). The frequency of MAIT cells co-expressing CCR6, the receptor for the CCL20 chemokine (a tissue-migrating marker) and survival CD127 marker was decreased, suggesting that the low circulating MAIT cell frequency might reflect their migration into inflamed infected tissues and/or activation-induced cell death (**Supplementary Fig. 3a**). MAIT cell activation in some patients might be associated with secondary infections and presence of bacteria in the blood (**Supplementary Fig. 3b**).

In contrast to MAIT cells, CD69^+^ expression on CD8^+^ T cell remained moderate (median of 10%) in both IDU and ICU although increased compared to uninfected controls (**Supplementary Fig. 3c**). Frequency of CD56^+^ CD8^+^ T cells was similar in all three groups of patients. Consequently, the frequency of effector CD56^+^ CD69^+^ CD8^+^ T cells was modestly increased in COVID-19 patients compared to uninfected controls (**Supplementary Fig. 3c**).

**Figure 2.**
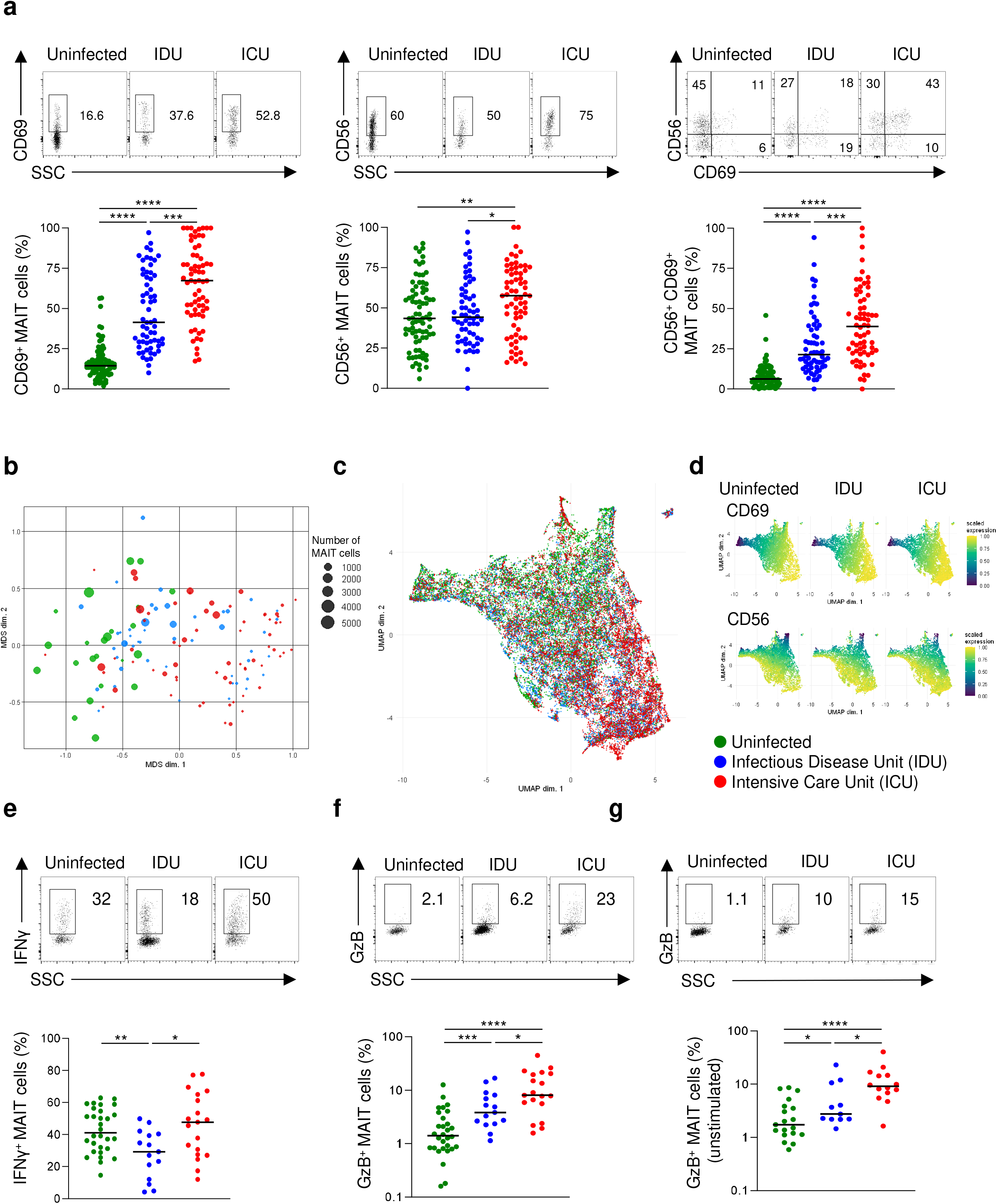
Blood MAIT cells are activated and secrete pro-inflammatory cytokines in COVID-19 patients. (**a**) Representative dot plots of CD69, CD56, and double-positive surface markers expression on blood MAIT cells from one uninfected control and two COVID-19 patients admitted in Infectious Disease Unit (IDU) and Intensive Care Unit (ICU) respectively. CD69^+^, CD56^+^, and CD56^+^ CD69^+^ MAIT cells frequencies in the blood of uninfected (n=80), and infected patients from IDU (n=62) and ICU (n=66). (**b-c**) Multi-Dimensional Scaling (MDS) plot (**b**) and Uniform Manifold Approximation and Projection (UMAP) (c) analysis of MAIT cells for each uninfected (n=23) and infected patient from IDU (n=50) or ICU (n=66) from Bichat hospital analyzed with the same flow cytometer. (**d**) UMAP divided by groups and colored by the scaled expression of CD69 and CD56. (**e-f**) Representative dot plots and frequencies of IFNγ (**e**), granzyme B (GzB) (**f**) in MAIT cells after stimulation from uninfected controls (n=25-27), COVID-19 patients hospitalized in an Infectious Disease Unit (IDU) (n=14-15) or hospitalized in an Intensive Care Unit (ICU) (n=15). (**g**) Representative dot plots and frequencies of GzB^+^ MAIT cells without stimulation in uninfected controls (n=19), IDU (n=11), and ICU (n=14) COVID-19 patients. SSC, side scatter. Small horizontal lines indicate the median, each symbol represents one biological sample. *P<0.05, **P<0.01, P<0.001, and *P<0.0001 (two-sided Mann-Whitney nonparametric test).

We next investigated with unsupervised methods the phenotype of MAIT cells, on 9 flow-cytometry parameters. We first compared MAIT cells of COVID-19 patients to uninfected controls by Multidimensional scaling (MDS) plots, which revealed a severity progression from non-infected controls to IDU and ICU patients (**Fig. 2b**). We next performed graphical dimensional reduction by both t-distributed Stochastic Neighbor Embedding (t-SNE) and Uniform Manifold Approximation and Projection (UMAP) that also returned a progressive distribution of MAIT cells from controls to IDU and ICU patients (**Fig. 2c** and **Supplementary Fig. 4a,b)**. Of note, both CD69 and CD56 markers were critical molecules in the differential distribution (**Fig. 2d** and **Supplementary Fig. 4c**).

MAIT cell function was then assessed after PMA/ionomycin stimulation by analyzing IFNγ and GzB production in all three groups of patients and IL-2, IL-4, IL-10, IL-17 and TNF in IDU and ICU patients (**Fig. 2e,f** and **Supplementary Fig. 5a**). Production of IFNγ was decreased in COVID-19 IDU patients compared to non-infected controls. However, IFNγ and IL-2 increased in ICU patients compared to IDU patients (**Fig. 2e** and **Supplementary Fig. 5a**). In contrast, GzB production progressively increased in stimulated MAIT cells in IDU and ICU patients as compared to controls. Elevated GzB production was also detected in unstimulated MAIT cells from infected patients (**Fig. 2f,g** and **Supplementary Fig. 5b**). Such enhanced cytokine and GzB production in ICU patients were not observed in conventional αβT cells, γδT cells and CD3^-^ cells (including NK cells) (**Supplementary Fig. 5a,b)**. Altogether, these results show that blood MAIT cells from COVID-19 patients display an activated/effector phenotype and cytotoxic function associated with disease severity.

### Links between MAIT cell activation and other innate immune cell alterations

Following MAIT cell analyses, we further investigated other innate-like immune cell frequency, phenotype and activation in the blood of SARS-CoV-2 infected patients. A significant NK cell, ILC2 and ILC3 frequency reduction in IDU and ICU patients was observed, which was more pronounced for NK cells and ILC2 in COVID-19 patients from ICU (**Fig. 3a**). CD69 expression was higher in all infected patients on NK cells, ILC3, and γδT cells whereas it was reduced on ILC2 (**Fig. 3b**).

**Figure 3:**
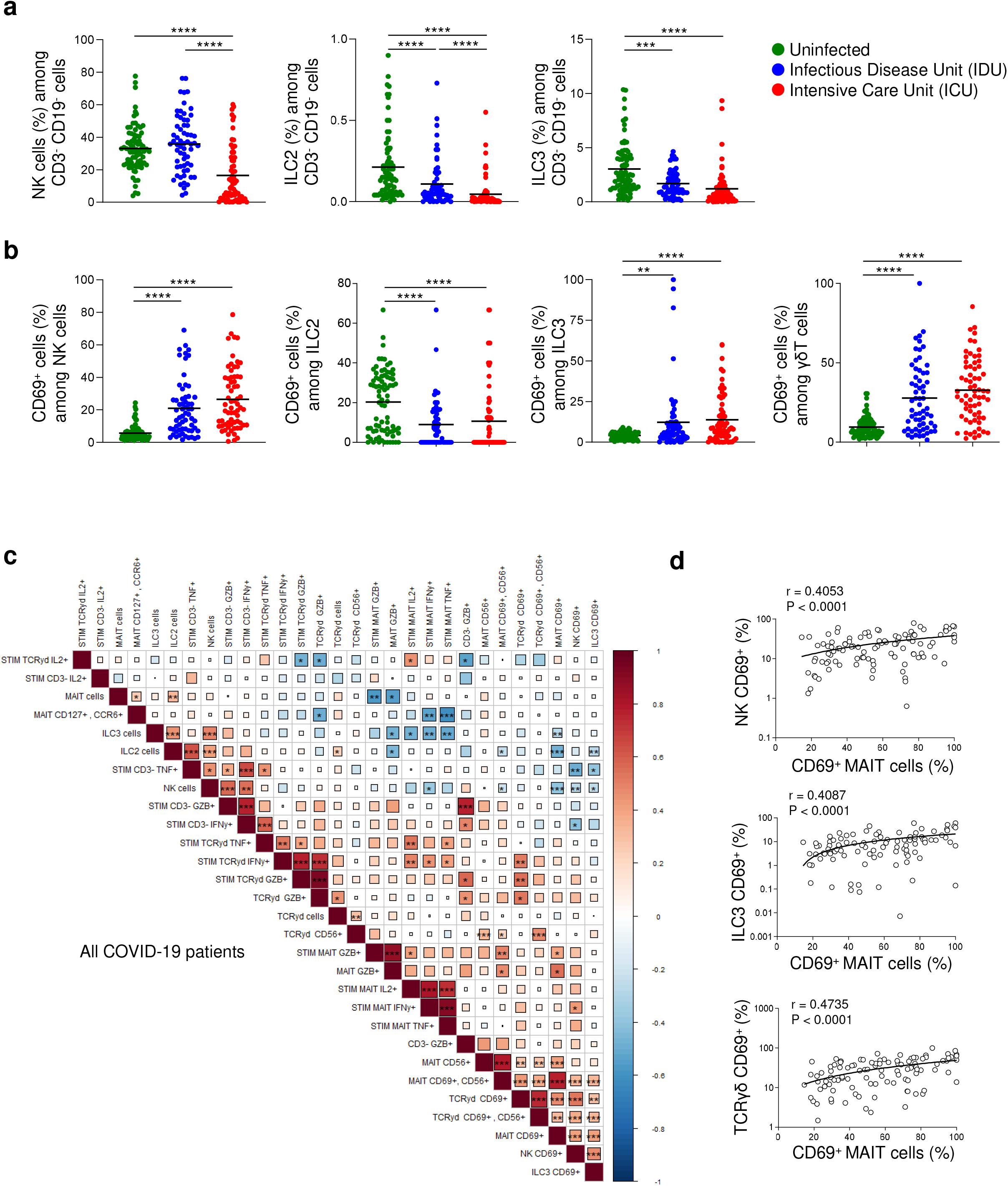
MAIT cells phenotype correlates with ILC and γ5T cells alteration in COVID-19 patients. **(a-b)** Frequencies of NK, ILC2, and ILC3 innate immune cells among CD3^-^ CD19^-^ cells **(a)** and CD69 activation surface marker on NK, ILC2, ILC3, and γδT cells **(b)** in uninfected controls (n=80), COVID-19 patients hospitalized in an Infectious Disease Unit (IDU) (n=62) or hospitalized in an Intensive Care Unit (ICU) (n=66). **(c)** Multiparametric matrix correlation plot of MAIT, γδT, ILCs, and NK cells frequencies, surface marker, and intracytoplasmic staining in COVID-19 patients. Spearman’s correlation coefficients are visualized by square size and color intensity. Variables are ordered by hierarchical clustering. **(d)** Correlation between CD69^+^ MAIT cells and CD69^+^ NK, CD69^+^ ILC3 or CD69^+^ γδT cells in COVID-19 patients (n=102). Small horizontal lines indicate the median, each symbol represents one biological sample. *P<0.05, **P<0.01, P<0.001, and *P<0.0001 (two-sided Mann-Whitney nonparametric test and Spearman nonparametric correlation test corrected for multiple inferences using Holm’s method).

We sought to identify correlations between MAIT cells and other innate cells in COVID-19 patients. A multiparametric matrix correlation plot showed strong positive correlations between the frequencies of activated MAIT cells (both CD56^+^ and/or CD69^+^ MAIT cells) with CD69^+^ ILC3, NK and γδT cells frequencies as well as CD69^+^ CD56^+^ γδT cells (**Fig. 3c,d)**. Several negative correlations were also observed between activation of these populations and their frequencies. CD69 expression on MAIT cells was negatively correlated with ILC3, ILC2 and NK cell frequencies (**Fig. 3c**). Moreover, cytokine production by MAIT cells was also negatively correlated with ILC3 frequencies and to a lesser extent with ILC2 and NK cell frequencies. Taken together, these data suggest that inflammatory processes in SARS-CoV-2-infected patients involve concomitant activation of MAIT cells with other innate-immune cells associated with loss of these populations’ frequencies in blood.

### Fatal SARS-CoV-2 infection is linked with activation and function of MAIT cells

To investigate the impact of immune cell populations on disease outcome, blood samples were analyzed by comparing two groups, surviving versus fatal outcome of COVID-19 infected patients. CD69 expression significantly increased on MAIT, CD8 T, γδT, and NK cells in fatal COVID-19 patients compared to surviving IDU and/or ICU patients (**Fig. 4a**). MAIT cells displayed the highest activation level in all patients, particularly those with fatal outcomes. We next examined immune cell function relative to disease outcome by measuring intracellular cytokines and GzB in stimulated and unstimulated immune cells (**Fig. 4b**, **Supplementary Fig. 6a,b** and data not shown). IFNγ, TNF, and GzB in stimulated MAIT cells and GzB in unstimulated MAIT cells were significantly enhanced in deceased compared to surviving patients with SARS-CoV-2 infection (**Fig. 4b** and **Supplementary Fig. 6a,b)**. IL-10 production by stimulated γδT cells in patients who succumbed was greater compared to surviving COVID-19 patients (**Fig. 4b** and **Supplementary Fig. 6a**). Thus, MAIT cell function was more associated to mortality than functions of other T cell populations. No modification of IL-2, IL-4, and IL-17 production in CD4^+^, CD8^+^, CD3^-^, γδT, and MAIT cells was observed in fatal cases (**Fig. 4b** and **Supplementary Fig. 6a**). Moreover, statistical regression analyses identified four MAIT cell markers (IFNγ production, stimulated GzB production, unstimulated GzB production, CD69) that defined predictive models for COVID-19 outcome, as tested on a receiver-operating-characteristic (ROC) curve (**Fig. 4c**). In order to have a more global view of individual patients, we generated an heatmap based on multiple immune cell and blood parameters from single uninfected, infected and fatal COVID-19 patients. This analysis highlights activation signatures of non-surviving patients compared to surviving COVID-19 patients and controls (**Fig. 4d**).

**Figure 4.**
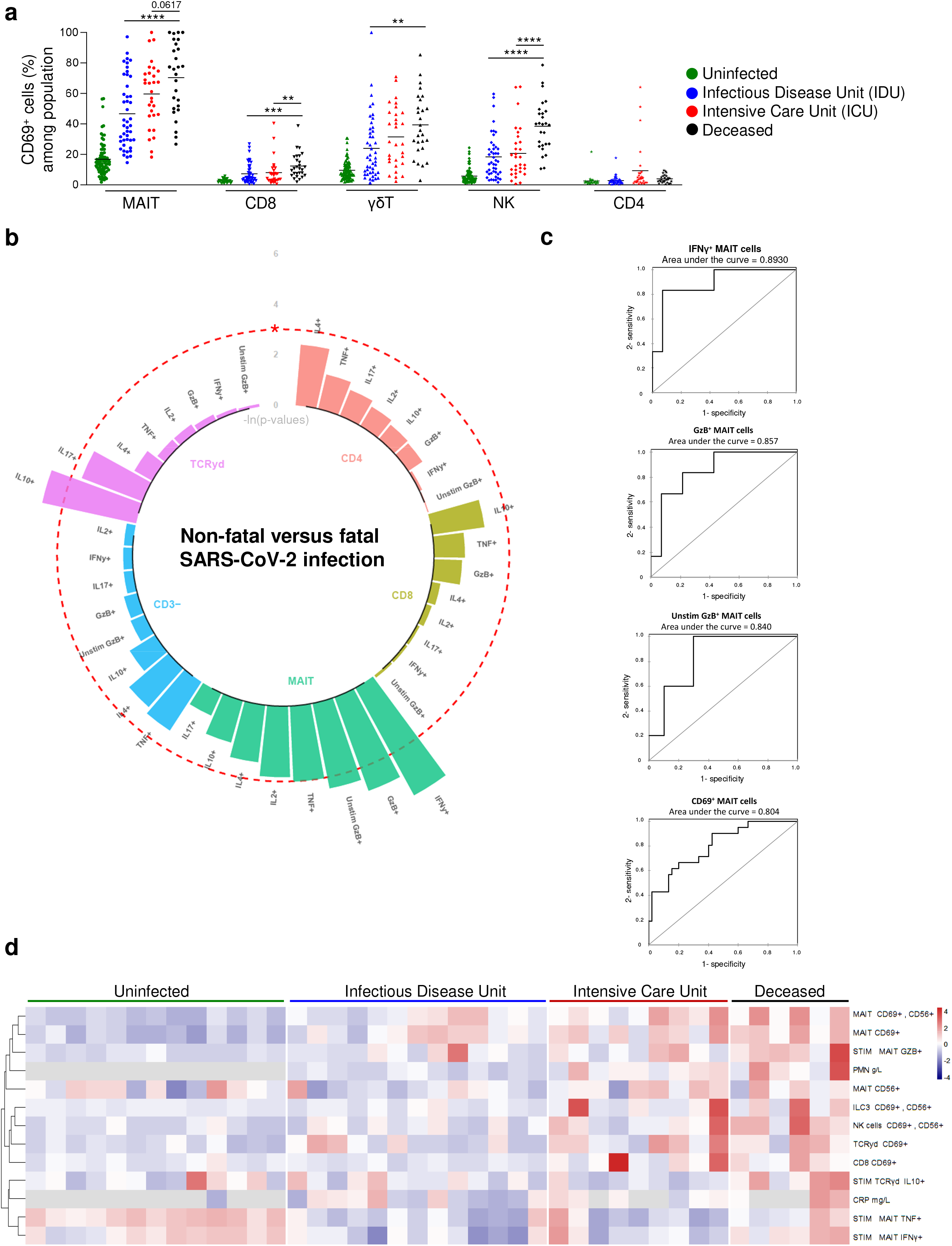
MAIT cell activation and functions are altered in fatal compared to non-fatal COVID-19. **(a)** Frequency of CD69^+^ cells among MAIT cells, conventional CD8^+^ and CD4^+^ T cells, γδT and NK cells in the blood of uninfected controls (n=80), surviving COVID-19 cases hospitalized in an Infectious Disease Unit (IDU) (n=45) or hospitalized in an Intensive Care Unit (ICU) (n=30) and fatal cases from both IDU and ICU (n=27). **(b)** Circular barplot representative negative natural logarithm p-values of the frequency of immune cells (MAIT, CD8^+^, CD4^+^, CD3^-^, γδT) function including IL-2^+^, IL-4^+^, IL-10^+^, IL-17^+^, IFNγ^+^, TNF^+^, and GzB^+^ after stimulation or GzB^+^ without stimulation (Unstim) between fatal (n=5-6) and non-fatal (n=16-23) SARS-CoV-2 infected patients. The red-dotted circle represents a p-value of 0.05. **(c)** Receiver-operating-characteristic (ROC) curves of the predictive MAIT cell markers defining the outcome of COVID-19. **(d)** Heatmap showing the scaled expression of different innate-like immune cells activation (MAIT, ILC3, NK, γδT, CD8), C-reactive protein (CRP), and polynuclear neutrophil (PPN) ordered by hierarchical clustering in uninfected controls (n=13), IDU (n=13), non-fatal ICU (n=9) and fatal ICU (n=6). Each symbol **(a)** or columns **(d)** represents one patient. Up-regulated parameters are shown in red, and down-regulated parameters are shown in navy blue. *P<0.05, **P<0.01, P<0.001, and *P<0.0001 (two-sided Mann-Whitney nonparametric test).

### Increased pro-inflammatory cytokine levels correlate with blood MAIT cell alterations

We next analyzed plasma levels of several cytokines by Cytometric Bead Array (CBA) and MSD Quickplex in both surviving IDU and ICU patients as well as non-surviving patients (**Fig. 5a,b)**. IL-6, IL-8, IL-10, IL-15, and IL-18 levels were significantly increased in the plasma of non-surviving patients compared to surviving patients, confirming a state of widespread, pronounced inflammation in severe COVID-19 cases ^28^ (**Fig. 5a,b)**. FNα2 levels were significantly decreased in ICU compared to IDU, although no significant difference between surviving ICU and deceased patients was detected (**Fig. 5b**). IL-1β levels were similar between all three groups (**Fig. 5b**). Multiparametric matrix correlation plot showed strong positive correlation of IL-6, IL-8, IL-10, IL-15, IL-18 levels with frequencies of CD69^+^, and CD69^+^ CD56^+^ MAIT cells in the blood of all COVID-19 patients (**Supplementary Fig. 7a,b)**. IFNα2 blood level correlated positively with the production of IFNγ, TNF, and IL-2 by MAIT cells (**Supplementary Fig. 7a**).

**Figure 5.**
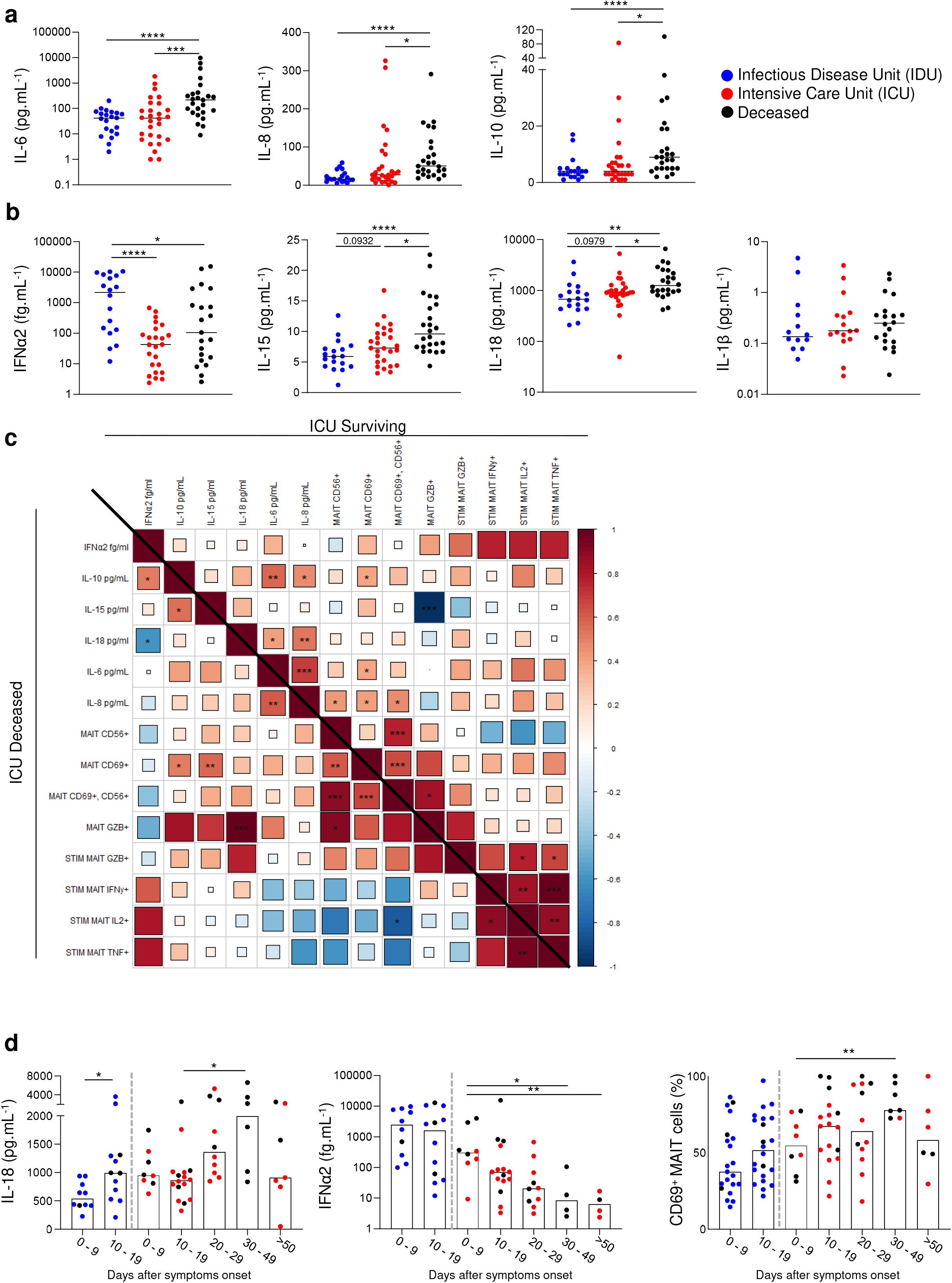
Pro-inflammatory cytokines are elevated in COVID-19 patients and match with MAIT cell activation and cytokine secretion. **(a)** Cytometric Bead Array (CBA) cytokine quantification of IL-6, IL-8, and IL-10 in the blood of surviving COVID-19 cases hospitalized in Infectious Disease Unit (IDU) (n=19-22) or hospitalized in Intensive Care Unit (ICU) (n=19-28) and fatal cases (n=25). **(b)** MSD Quickplex cytokine quantification of IFNα2, IL-1β, IL-15, and IL-18, in the blood of surviving COVID-19 cases hospitalized in IDU (n=14-22) or hospitalized in ICU (n=16-28) and fatal cases (n=20-25). Values under the limit of detection are not statistically computed and are not displayed. **(c)** Multiparametric matrix correlation plot of IL-6, IL-8, IL-15, IL-18, IFNα cytokines blood level; frequencies of CD69^+^, CD56^+^, CD69^+^ CD56^+^, and GzB^+^ MAIT cells; frequencies of IFNγ^+^, IL-2^+^, TNF^+^, and GzB^+^ stimulated MAIT cells; in non-fatal (upper right part, n=30) versus fatal COVID-19 patients in ICU (lower left part, n=21). Spearman’s correlation coefficients are visualized by square size and color intensity. Variables are ordered by alphabetical order. **(d)** CD69^+^ MAIT cells, IL-18, and IFNα blood levels according to symptoms duration (days) in COVID-19 patients, starting at the first clinical sign. Small horizontal lines indicate the median, each symbol represents one patient. *P<0.05, **P<0.01, P<0.001, and *P<0.001 (two-sided Mann-Whitney nonparametric test and Spearman nonparametric correlation test corrected for multiple inferences using Holm’s method).

Separated matrix correlation plot between surviving and non-surviving ICU patients showed different relationships between blood cytokines and GzB production by MAIT cells. In surviving patients, there was a strong negative correlation between blood IL-15 level and spontaneous GzB production by MAIT cells whereas in non-surviving patients there was a strong positive correlation between blood IL-18 level and GzB MAIT production. The link between IL-18 and MAIT cell activation is further supported by high IL-18Rα expression on all blood MAIT cells in control and COVID-19 infected compared to other immune populations (**Supplementary Fig. 8a,b,c)**. Interestingly, in non-surviving ICU patients, blood IL-18 level was negatively correlated with IFNα2 blood levels (**Fig. 5c**). Accordingly, IL-18 blood level was increased in long-term infected ICU patients whereas IFNα2 blood level was decreased in long-term in these patients (**Fig. 5d**). Of note, CD69^+^ MAIT cell frequency increased as well with time and was highest one month after symptoms onset, when death rate was highest among ICU patients. Our data therefore reveal that pro-inflammatory cytokines and IL-10 levels are associated with MAIT cell activation and highlights a unique relationship between plasma IL-18 levels and circulating cytotoxic MAIT cells in fatal cases.

### MAIT cell phenotype and functions are associated with SARS-CoV-2 severity and clinical parameters

As previously reported in our cohort, COVID-19 severity was associated with extended pulmonary damage as evaluated on chest computed tomography (CT) (score from 1:mild to 5:critical) (**Fig. 6a**). PaO_2_/FiO_2_ ratio was significantly reduced and C-reactive protein (CRP) levels were significantly increased in ICU patients. Both parameters were even more impacted in fatal cases (**Fig. 6b,c)**.

**Figure 6:**
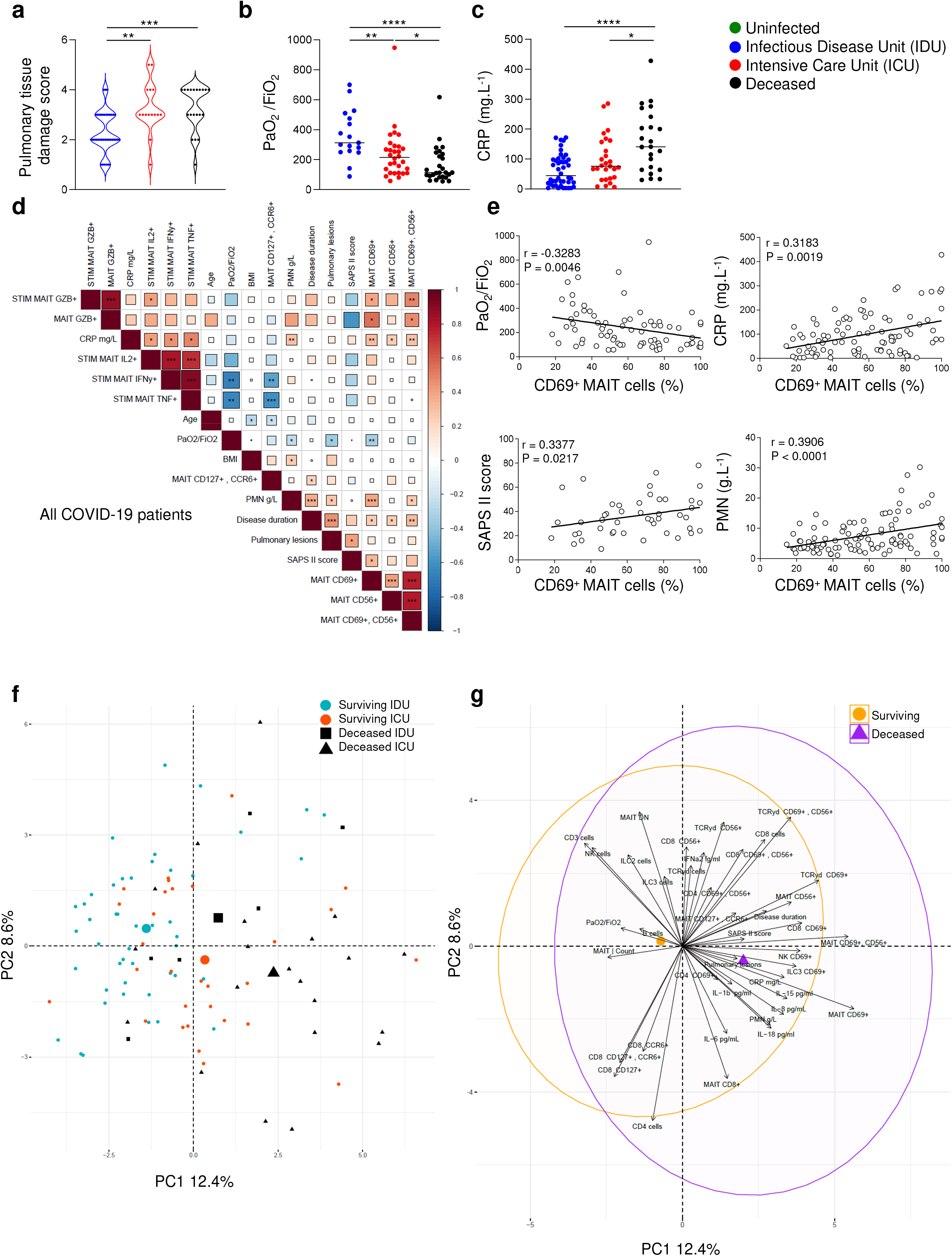
MAIT cell phenotype and blood cytokine levels are associated with SARS-CoV-2 infection severity. **(a-c)** Pulmonary damages score level (1: light - 5: critical) **(a)**, PaO_2_/FiO_2_ ratio **(b)** and C-Reactive protein (CRP) **(c)** of surviving COVID-19 cases hospitalized in Infectious Disease Unit (IDU) (n=38, n=17, n=44) or hospitalized in Intensive Care Unit (ICU) (n=15, n=30, n=27) and fatal cases from IDU and ICU (n=20, n=27, n=23). **(d)** Multiparametric matrix correlation plot of the following clinical data: age, BMI, CRP, SAPS II score, PaO_2_/FiO_2_, Polymorphonuclear neutrophils (PMN), Disease duration, Pulmonary lesions score); frequencies of CD69^+^, CD56^+^, CD69^+^ CD56^+^, CCR6^+^ CD127^+^, and GzB^+^ MAIT cells; frequencies of IFNγ^+^, IL-2^+^, TNF^+^, and GzB^+^ stimulated MAIT cells; in the blood of all COVID-19 patients. Spearman’s correlation coefficients are visualized by square size and color intensity. Variables are sorted by hierarchical clustering. **(e)** Correlation between PaO_2_/FiO_2_ ratio (n=73), SAPS II score (n=46), CRP (n=94), and PMN (n=101) and the frequency of CD69^+^ MAIT cells in blood (presented as a % value of total MAIT cells) from all COVID-19 patients. **(f-g)** Principal component analysis (PCA) of 50 variables (listed in **Supplementary Fig. 9**) including clinical data and frequencies of immune cell phenotype in IDU (n=46), ICU (n=30), and fatal cases from IDU (n=6) and ICU (n=21) infected patients. Each point represents a single patient. Mean value of each group is symbolized by a bigger symbol **(f)**. Arrows represent quantitatively the contributions of each variable in the PCA, the first 40 contributing parameters are displayed. Mean values of groups are represented by a symbol. Concentration ellipses with a confidence of 95% are shown **(g)**. Small horizontal lines indicate the median, each symbol represents one patient. *P<0.05, **P<0.01, P<0.001, and *P<0.0001 (two-sided Mann-Whitney nonparametric test and Spearman nonparametric correlation test corrected for multiple inferences using Holm’s method).

A correlation matrix of MAIT cells activation, function and clinical parameters was established for all COVID-19 patients (**Fig. 6d**). It included age, body mass index (BMI), CRP, Simplified Acute Physiology Score II (SAPSII) (an estimator of patient mortality risk at ICU admission), PaO_2_/FiO_2_ ratio, polymorphonuclear neutrophil (PMN), disease duration and pulmonary lesions. Analysis of these clinical indicators in light of MAIT cells revealed that on vital parameters, PaO2/FiO2 strongly negatively correlated with CD69^+^ expression, IFNγ and TNF production by MAIT cells, whereas SAPSII score positively correlated with CD69^+^ MAIT cell frequency (**Fig. 6d,e)**. Concerning inflammatory markers, CRP level correlated with MAIT cell activation phenotype and cytokine secretion (IL-2, IFNγ and TNF) while PMN frequency positively correlated with CD69^+^ MAIT cell frequency (**Fig. 6d,e)**.

Principal Component Analysis (PCA) of all COVID-19 patients showed an unsupervised overview of all clinical data, cytokine levels, immune cell frequencies, activation, and functions for each COVID-19 patient (**Fig. 6f**). The PCA showed the segregation of deceased patients compared to surviving patients with CD69^+^ MAIT cells being the most contributing variable (**Fig. 6g** and **Supplementary Fig. 9**). Of note, several parameters also contributed to the surviving/fatal cases discrimination vector such as blood IL-8, IL-15, IL-18, CRP, PMN, activated ILC3 and NK cells that we previously found to be correlated to MAIT cell activation (**Fig. 3c,d, Supplementary Fig. 9**, and **Fig. 6d,e)**. Therefore, altered activation markers and cytokine production by blood MAIT cells correlate with clinical parameters and are associated with both severity and disease outcome.

### SARS-CoV-2-infected macrophages activate MAIT cell *in vitro*

MAIT cells exert antiviral properties that are promoted especially by macrophages ^18,20^. It has been established that macrophages and monocytes functions are impacted by SAR-CoV-2 infection^26,27^. Using an *in vitro* co culture model we analyzed whether SARS-CoV-2-infected macrophages impacted MAIT cell phenotype. Blood monocytes isolated from healthy donors were cultured during 7 days and differentiated into mature macrophages (GM-MΦ). GM-MФ were then infected with SARS-CoV-2 at different multiplicity of infection (MOI) (0.3 or 3) during 2 hours and then cocultured with autologous PBMC or enriched MAIT cells for 24 to 96 hours (**Fig. 7a**). Infected macrophages of 4 healthy donors (MOI=0.3 or 3) induced increased expression of CD69 on MAIT cells, when compared to uninfected macrophages (mock), in a dose dependent manner (**Fig. 7b**). Of note, although macrophage infection by SARS-CoV-2 was similar in all donors (**Fig. 7c**), level of MAIT cell activation by infected macrophages varied, suggesting intrinsic differences between individuals that modulate MAIT cell response upon infection. Other innate immune cells (γδT and NK cells) were also activated by infected macrophages, however, to a lesser extent than MAIT cells. CD8, CD4 T cells or ILC3 co-culture (MOI=3) were almost unaffected (**Fig. 7d**). Analysis of several donors showed that MAIT cells were always activated, whereas it was not the case for other innate immune cells (**Fig. 7e**).

**Figure 7:**
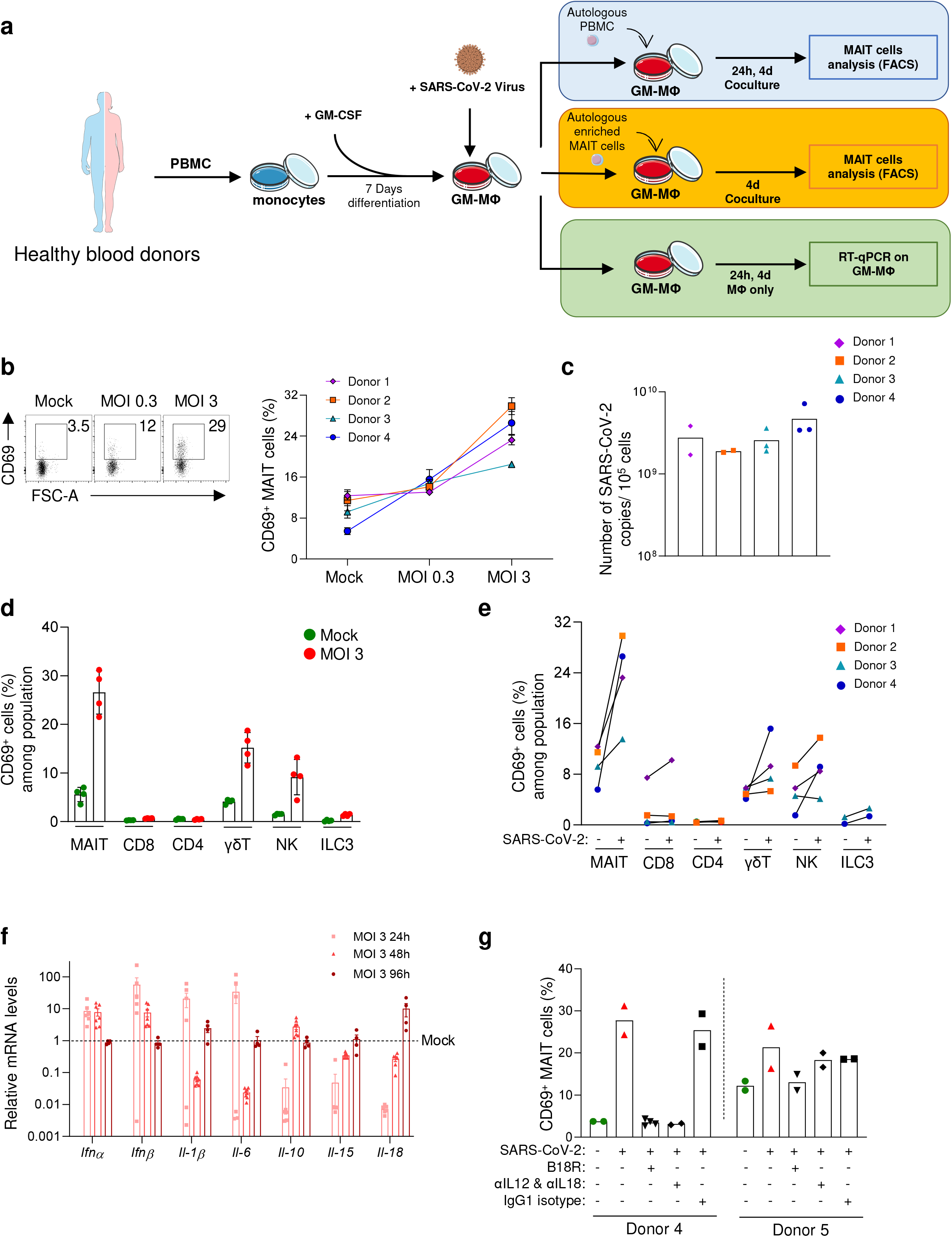
Infected macrophages are altered and trigger MAIT cells *in vitro*. PBMCs of uninfected patients are challenged with infected macrophages with the SARS-CoV-2 virus. **(a)** Graphical flowchart of the *in-vitro* co-culture experiments. **(b)** Representative dot plot and frequency of CD69^+^ MAIT cells in 4 healthy donors co-cultured with different SARS-CoV-2 Multiplicity of Infection (MOI) with mock, 0.3, and 3, with replicates (n=2-4). Each symbol represents the mean value of replicates. **(c)** Viral quantification of SARS-CoV-2 in co-culture from four donors at 96h, by RT-qPCR. **(d-e)** Frequency of CD69^+^ cells among MAIT cells, conventional CD8^+^ and CD4^+^ T cells, γδT and NK cells in donor n°4 **(d)** and all donors **(e)** co-culture at 96h, with replicates (n=2-4). **(f)** Kinetic of *Ifnα, Ifnβ, Il-1β, Il-6, Il-7, II-10, II-15*, and *Il-18* mRNA relative levels in infected macrophages from two healthy donors at 24h, 48h, and 96h after infection. **(g)** Co-culture of infected macrophages (MOI 3) with PBMC (donor 4) or purified T cells (donor 5) with or without anti-IL-12, anti-IL-18, or recombinant soluble IFNα receptor protein (B18R), with replicates (n=2-4). Each symbol represents a single co-culture well.

Since MAIT cells exert their antiviral activity in a cytokine dependent manner^18,20^, we next analyzed cytokine production by infected macrophages. Gene expression analysis showed that a short-term infection of macrophages by SARS-CoV-2 (24 hours) induced upregulation of their *Ifnα, Ifnβ, Il-1β* and *Il-6* expression compared to uninfected controls whereas *Il-10, Il-15* and *II-18* expression was downregulated. While *Ifn*α, *Ifnβ, Il-1β, Il-6, II-10* and *II-15* expression return to basal levels 4 days after infection, the expression of *Il-18* was upregulated (**Fig. 7f**). Finally, we observed that blockade of type I IFN, IL-12 and IL-18 completely inhibited MAIT cell activation of the donor 4 whereas it was only blocked by type I IFN inhibitor for the donor 5 (**Fig. 7g**). Altogether this data suggests that MAIT cell activation upon infection by SARS-CoV-2 is a two-step process. Early infection may induce MAIT cell activation through type I IFN pathways and later through the IL-18 pathway.

## Discussion

Our study reveals major MAIT cells alteration in numerous COVID-19 cases. MAIT cell frequency is strongly reduced in greater proportion than all other major T cell subsets and they are highly activated with secretion of critical pro-inflammatory cytokines, such as IFNγ. MAIT cells possess a strong cytotoxic capability with an increased expression of GzB. Moreover, all these MAIT cell alterations scale with the severity of SARS-CoV-2 infection, from mild to fatal cases, and correlate with both plasmatic pro-inflammatory cytokine levels and other innate cell activation. *In vitro* experiments identified a SARS-CoV-2 macrophage cytokine shift and their ability to stimulate MAIT cells.

The frequency of blood MAIT cells was extremely reduced, down to a tenth compared to controls in severe cases of COVID-19^30^. Several hypotheses might explain this loss of blood MAIT cells. The tissue migration chemokine receptor CCR6 expression is decreased on blood MAIT cells during SARS-CoV-2 infection whereas its ligand, CCL20, is expressed by pro-inflammatory pulmonary macrophages ^31^. Collapse of MAIT cell frequency in blood may be due to migration of CCR6^+^ MAIT cells in the infected lung where they might participate to local immune response, as already demonstrated in other pulmonary infections mediated both by bacteria or viruses ^18,19^. In addition, a recent study in a limited number of COVID-19 patients showed an increased pulmonary MAIT cell frequency in the broncho-alveolar lavage fluid ^30^. Low blood MAIT cell frequency may also be due to their apoptosis. Expression of CD127, receptor for the pro-survival cytokine IL-7, is reduced on blood MAIT cells from infected patients. Moreover, MAIT cells are terminal effector T cell with high GzB expression, suggesting limited lifespan^32^.

MAIT cell frequency and activation alterations are characteristic of several deleterious inflammatory pathologies, especially metabolic pathologies such as T2D, liver disease or obesity^10,33-36^. MAIT cell frequency is reduced in blood from patients with those pathologies and they present an exhausted phenotype that may impair their antibacterial response^34-36^. This is of importance as metabolic pathologies are important factors increasing risk of developing severe COVID-19^37-40^. Thus, in our study, we matched our controls for comorbidities affecting COVID-19 patients, mainly obesity and diabetes. More than half of the ICU patients of our cohort developed secondary infections from mild tissue-specific infections to life-threatening sceptic shocks. Previous studies showed that high frequency of MAIT cells is associated with survival during sceptic shocks^41^. Similarly, patients co-infected by *Mycobacterium tuberculosis* and HIV are more vulnerable to tuberculosis because of a reduced MAIT cell frequency in blood^23,42^. COVID-19 patients with metabolic syndromes may therefore be extremely vulnerable to complications because of a diminished ability to fight both primary viral infection and secondary bacterial infections to a persistent depletion of blood MAIT cells.

Our data highlights strong positive correlations between blood NK, ILC3, γδT and MAIT cell activation. In other lung pathologies such as asthma, CD69^+^-activated NK, ILCs, and MAIT cells are associated with airflow limitation^43^. We observed a significant correlation between MAIT cells activation and ILC2 frequency and it is already established that ILC2 are important drivers of allergen-induced airway hyperresponsiveness (AHR) after influenza A virus (IAV) infection^14^.

Blood cytokine analysis showed increased amounts of type I interferons and pro-inflammatory cytokines such as IL-6, IL-8 and the immunosuppressive IL-10 cytokines in COVID-19 patients as previously reported^28^. Presence of elevated levels of IL-1β, IL-6, IL-8, and IL-10 are indicative of cytokine storms that are major potential complications in severe COVID-19 patients. Moreover, IL-15 and IL-18 levels were increased in COVID-19 patients in relation with disease severity and IL-18 strongly correlated with MAIT cell activation in patients with fatal outcome. In these patients, IFNα2 level inversely correlated with increased IL-18. During the early phases of infection *in vitro*, macrophages produced important amounts of type I interferons which collapsed in later stages, around 4 days post-infection. This is in agreement with the ability of SARS-CoV-2 to suppress type I interferon production through numerous structural and non-structural viral proteins^44^ and as highlighted in COVID-19 patient plasma analysis of ICU patients. However, long-term *in vitro* infected macrophages produce increased levels of IL-18 that can activate MAIT cells in COVID-19 corroborating the extremely high blood IL-18 level correlating with MAIT cell activation in deceased patients. Both IL-15 and IL-18 are the most effective pro-inflammatory cytokines capable to activate MAIT cells in a TCR-independently manner during viral infection^19^. CD56^+^ MAIT cells, which are increased in severe cases from our COVID-19 cohort, are known to have a higher capacity to respond to type I IFN and IL-18 cytokines that are key players in MR1-independent MAIT cell responses during viral infection^45^.

MAIT cells in inflammatory pathologies are double-edged swords. They can be protective by participating in and supporting pathogen clearance, immune activation and tissue reparation^10,12,46^. Patients infected with pandemic IAV harbor an inverse correlation between MAIT cell frequencies and disease severity, suggesting a protective role for MAIT cells^18,20^. Conversely, MAIT cells may fuel detrimental inflammation. Our data suggests a negative role for MAIT cells in severe COVID-19 infection in which their activation and GzB production are the highest. Against infection with Dengue virus (DV), there is a temporal and quantitative association between the activation of MAIT cells and the onset of severe disease^20^. It is interesting to note that both the DV and the SARS-CoV-2 are able to infect macrophages, which then can activate MAIT cells^47^. Activation of MAIT cells by infected macrophages through IL-18 may switch MAIT cells toward a detrimental role in these infections.

In conclusion, human MAIT cells are activated, displaying a cytotoxic profile in the blood of SARS-CoV-2 infected patients, which is associated with other innate immune cell activation, and a pro-inflammatory environment. Together, these data extend the knowledge of the immune actors involved during SARS-CoV-2 infection. These findings reveal MAIT cells as a valuable biomarker of disease progression and a new target for interventional therapeutic approaches in severe SARS-CoV-2 infection.

## Data Availability

Data and code generated during this study will be available before publication in a repository. Access code will be given upon request at

## Materials and methods

### Clinical study design and ethical statement

One-hundred and two COVID-19 patients admitted in Bichat or Cochin hospitals, Paris, France, between March 23, 2020 and May 29, 2020 were included in this clinical study. For comparisons, blood from 80 uninfected controls of the Quid-Nash project (n=30), Etablissement Français du Sang (EFS) (n= 12), and volunteer donors (n=38) were mostly collected before pandemic onset (54/80). Clinical characteristics of the 182 patients are summarized in Fig. 1a and further detailed in Supplementary Table 1. No statistical methods were used to predetermine cohort size. The percentage of lung involvement was evaluated on chest CT with a score of 1 to 5 to represent mild to critical pulmonary lesions damages. Simplified Acute Physiology Score (SAPS II) was used to estimate the probability of survival after Intensive Care Unit (ICU) admission. The Ethics Committees approved clinical investigations. Informed consent was obtained from each enrolled patient. Patient’s from Bichat Hospital (Paris, France) were included in the French COVID cohort (NCT04262921). Ethics approval for this cohort was given on February 5th, 2020 by the French Ethics Committee CPP-Ile de France-VI (ID CRB: 2020-A00256-33). This cohort is sponsored by Inserm and supported by the REACTing consortium and by the French Ministry of Health (PHRC n°20-0424). Samples from these patients were derived from samples collected in routine care. Patients from Cochin Hospital (Paris, France) were recruited in the setting of the local RADIPEM biological samples collection derived from samples collected in routine care. Biological collection and informed consent were approved by the Direction de la Recherche Clinique et Innovation (DRCI) and the French Ministry of Research (N°2019-3677). Investigations with control patients from QUID-NASH were approved by Comité de Protection des Personnes de Sud Méditerranée (V) #18.021, N° of QUID project registration: 2018-A00311-54.

### Whole blood, PBMC and plasma isolation

Whole blood samples were collected in EDTA or heparin-coated tubes (Vacutaine, BD Biosciences) from healthy, non-infected, donors; and COVID-19 patients admitted to Cochin, Bichat, or Lariboisière Hospitals in Paris, France. Tubes were centrifugated at 1260g for 10 minutes and plasma was collected and frozen at −80°C. PBMCs were isolated by Ficoll-Paque (Lymphosep, Biosera), frozen in 1mL of freezing medium constituted of 90% FCS and 10% DMSO and stored in liquid nitrogen.

### Cytokine measurements

IL-6, IL-8 and IL-10 were measured in plasma using Human Cytometric Bead Array (CBA) Inflammatory Cytokine Kit (BD Biosciences) according to manufacturer’s instructions. Acquisitions were performed on a BD FACSLyric cytometer (BD Biosciences) and raw data analyzed with FCAP Array software V3.0 (BD Biosciences). For the IFNα2 quantification, plasma samples were analyzed with the MSD Quickplex using the Ultra-sensitive assay S-PLEX Human IFN-α2a (reference K151P3S-1) from Meso Scale Diagnostic MSD (Rockville, US) using 25 μL of each sample. Each plasma sample was assayed twice with the average value taken as the final result. The unit for Human IFN-α2a measured in the present study is fg.mL^-1^.

For IL-18, IL-15 and IL-1β quantification, the plasma samples were analyzed with the MSD Quickplex using the U-plex Biomarker group 1 (human) Assay (K-15067L-1) from Meso Scale Diagnostic MSD, using 25 μL of each (1/2 diluted) sample. Each plasma sample was assayed twice with the average value taken as the final result. The unit for Human IL18, Il-15 and IL-1β measured in the present study is pg.mL^-1^.

### Flow cytometry

Surface and intracytoplasmic staining were performed on blood samples or PBMC with the following antibodies: CD3 (OKT3), CD19 (HIB19), CD4 (OKT4), CD8 (SK1), Vα7.2 (3C10), TCR-γδ (B1), CD161 (HP-3G10), CCR6 (G034E3), CD56 (HCD56), CD69 (FN50), CD127 (R34.34), CD218a (H44), IFNγ (4S.B3), TNF (Mab11),, IL-2 (MQ1-17H12), IL-4 (8D4-8), IL-10 (JES3-19F1), IL-17 (BL168), granzyme B (GB11). Full list is described in **Supplementary Table 1**. According to the amount of blood obtained from each patient, surface staining was always performed, and depending on number of cells, intracytoplasmic staining of cytokines and granzyme B were analyzed after stimulation with PMA-ionomycin-brefeldin A. Biotinylated human MR1 tetramers loaded with the active ligand 5-(2-oxopropylideneamino)-6-D-ribitylaminouracil (5-OP-RU) were used to confirm MAIT cells identification. MR1 tetramers were coupled to streptavidin-PE (National Institutes of Health tetramer core facility, USA).

For surface staining, staining of 200μL of blood sample was performed in PBS containing 1% BSA and 0.05% sodium azide. After surface staining, cells were fixated using BD FACS Lysing Solution (BD Biosciences, #349202) according to the manufacturer’s instructions. Data acquisition was performed using a BD Biosciences LSR Fortessa. Flow cytometric analyses were performed with the FlowJo analysis software V10.6.2 (Tree Star).

For intracellular labelling, thawed PBMCs were treated with DNAse (0.05 mg.mL^-1^) (#D4263, Sigma-Aldrich, USA) in RPMI (#61870-010, Gibco, USA) and incubated at 37°C, 5% CO_2_ during 30 minutes. Depending on the number of PBMCs obtained from each patient, intracytoplasmic staining of cytokines and granzyme B were analyzed with stimulation with PMA (#P-8139, Sigma-Aldrich, USA), ionomycin (#I-0634, Sigma-Aldrich, USA) and Brefeldin A (#B-7651, Sigma-Aldrich, USA) and/or only Brefeldin A. Stimulated cells were incubated during 6h at 37 °C in RPMI medium supplemented with 10% FCS, 1% HEPES, 1% Penicillin/Streptomycin and stimulated with PMA (25 ng.mL^-1^) and ionomycin (1 μg.mL^-1^), in the presence of brefeldin A (10 μg.mL^-1^). After surface staining, cells were fixed and permeabilized with a Cytofix/Cytoperm kit (#554714, BD Biosciences, USA), then were washed using Perm/Wash buffer (BD Biosciences, #554723) and incubated at 4 °C in the dark for 30 min with antibodies against cytokines and GzB (listed above).

### *In vitro* culture

For virus culture, the Vero E6 kidney epithelial cells line was acquired from the American Type Culture Collection (#CRL-1586, ATCC, USA) (LGC standards SARL, Illkirch, France). Vero E6 cells were cultured in DMEM (GibcoTM) supplemented with 10% of heat-inactivated fetal bovine serum (FBS, GibcoTM) (Thermo Fisher Scientific, Waltham, MA, 209 USA) and maintained at 37°C in a humidified atmosphere containing 5% CO_2_. The viral strain of human SARS-CoV-2 was obtained from a nasopharyngeal positive PCR sample. SARS-CoV-2 primo-culture stock used in this study was produced in Vero E6 cells and titrated by lysis plaque assay ^48^. SARS-CoV-2 stock titer was 2.10^7^ PFU.mL^-1^. Supernatant was aliquoted for storage at −80°C.

For viral titration, SARS-CoV-2 was titrated by lysis plaque assay as previously described ^49^. Vero E6 cells were plated onto 12-well plate at a density of 5.10^4^ cells per well in DMEM with 10% FBS. 24h later, cells were infected by 10 to 10 serial viral dilutions. After virus adsorption for 1h at 37°C with plate rocking every 15 min, the viral inoculum was removed and Vero cells were washed with PBS free medium. After, 500μL of an agarose medium mix was added. After three days of incubation at 37°C with 5% CO2, supernatant was removed and cells were fixed with 1 mL of 6% formalin solution for 30 minutes. The formalin solution was removed and cells were colored with a 10% crystal violet solution for 15 minutes. All wells were then washed with distilled water and dried on bench-coat paper before analysis.

For macrophages in vitro infection by SARS-CoV-2, CD14^+^ monocytes cells were isolated using StraightFrom® Whole Blood CD14 MicroBeads (#130-090-879, Miltenyi Biotec). 2.10^5^ monocytes from one donor were added per culture well (96-flat-well plate). Monocytes were differentiated into macrophages by incubating CD14^+^ monocytes cells for 7 days in X-VIVO15 (#BE02-060F, Lonza, Switzerland) with 50 ng.mL^-1^ of GM-CSF (#130-093-864, Miltenyi Biotec).

For infection, macrophages were washed two times in PBS and were treated with virus in non-supplemented RPMI at a MOI of 0.3 and 3 as determined based on the viral titer and the number of cells plated, for 90 to 120 min at 37 °C. The macrophages were washed three times with PBS before adding the lysis buffer into each well. The intracellular viral load Viral quantity was then determined by RT-qPCR (see below).

### PBMC preparation and coculture with macrophages

PBMC were isolated from fresh blood samples using Lymphocyte Separation Media lymphosep LM-T1702/500 (Biosera). B cells, monocytes, and CD4^+^ cells were depleted from PBMC of the same healthy donors using Dynabeads™ Untouched™ CD8^+^ kit (#11348D, Invitrogen, USA) with an in-house antibody mix with the following biotinylated monoclonal anti-human antibodies CD4 (OKT4), CD14 (63D3) and CD19 (HIB19) from Biolegend. 2.10^5^ PBMCs or enriched MAIT cells per well were co-cultured with differentiated macrophages during either 24h or 96 hours. Blocking antibodies were used against IL-12p70 (MAB219, R&D systems) at 5μg.mL^-1^ and IL-18 (D044-3, MBL) at 5μg.mL^-1^. 1 μg.mL^-1^ B18R (34-8185-81, eBioscience) was used to block Type I interferons^50^. After the coculture, macrophages were washed three times with PBS before adding the lysis buffer into each well. The intracellular viral quantity was then determined by RT-qPCR (see below).

### RNA extraction and RT-qPCR

Total nucleic acids from cells and supernatant were extracted with the Total NA Isolation kit - Large Volume assay on a MagNA Pure LC 2.0 analyzer (Roche). Nucleic acids were eluted in 50μL of elution buffer and immediately tested by a quantitative PCR of the albumin gene for cellular DNA^51^ and the RealStar™ SARS-CoV-2 RT-PCR Kit 1.0 (Altona Diagnostics GmbH) for SARS-CoV-2 detection. Viral RNA quantification was achieved using a standardized RNA transcript control acquired from the European Virus Archive Program.

### Cytokine RNA quantification

cDNA was produced from total extracted nucleic acids as described above using Superscript III reverse transcriptase (#18080044; Invitrogen). Primers sequences used are described in **Supplementary Table 2**. Quantitative PCR analysis was performed with SYBR Green (# 4887352001; Roche) and was analyzed with a LightCycler 480 (Roche). Relative expression was calculated by the 2^-ΔΔCt^ method and was normalized to expression of the housekeeping gene encoding 18S and, when detailed, normalized on mock-infected macrophages.

### Statistics and Bioinformatics analysis

All bioinformatics analyses were performed using RStudio (1.2.5) running on R software version 4.0. We used the flowcytometry analysis R workflow from Nowicka et. al^52^ to produce MDS (multidimensional scaling) plots of aggregated signal and normalized dimension reduction plot UMAP and t-SNE with 500 MAIT cells per patient from Bichat hospital flow cytometry data. Our pipeline combined this workflow with the flowWorkspace R library for FlowJo workspace reading and cell population selection. Multiparametric matrix correlation plots were produced with the adjusted rcorr function from Hmisc and RcmdrMisc packages to compute matrices of Spearman correlations along with the pairwise p-values corrected for multiple inferences using Holm’s method and visualized with the Corrplot package. Correlation plots hierarchical clustering were produced with the hclust function included in the Corrplot library. Observations were filtered for missing values, and only complete observations were used. Principal component analysis (PCA) was processed with FactoMineR library and graphically produced with Factoextra package. Heatmaps were plotted using pheatmap library, with data centered to zero and scaled for each parameter. Clinical table values were computed with the atable R library. All R code written and used from our study is available at github.com/MatthieuRouland/MAIT-COVID19.

Statistical analyses were performed with GraphPad Prism software version 8.0 and R software version 4.0. All datasets were tested for normal distribution using Shapiro-Wilk normality test. Since all normality tests returned negative, all datasets were compared using nonparametric two-tailed Mann-Whitney. Correlation calculation between two parameters has been performed using the Spearman’s correlation test corrected for multiple inferences using Holm’s method. Logistic regression and ROC (Receiver Operating Curve) were produced with XLSTATS 2020.4 and confirmed with a randomly-split cohort on R with the RORC package. Prognostic validity of the model was evaluated by analysis of the ROC curve and was measured using the area under the curve (AUC). Differences were considered significant at P < 0.05 (*P<0.05, **P<0.01, P<0.001, *P<0.0001).

## Acknowledgments

We thank all the patients and their physicians, nurses and technician staff who helped with the study. We thank the Department of Biological Hematology and the Department of Biochemistry of Bichat-Claude Bernard University Hospital for measuring white blood cells and C-reactive protein. We are grateful to Marc Diedisheim for biostatistics and bioinformatics discussion, Quentin Le Hingrat for help in virus experiments, Antoine Costa Monteiro for help in setting up the collaboration between virologists and immunologists, and the National Institutes of Health tetramer core facility for human MR1 tetramers. A.L. and R.C.M. laboratories are supported by ANR-11-IDEX-0005-02 Laboratory of Excellence INFLAMEX and Fondation pour la Recherche Médicale (EQU201903007779 to A.L. and EQU201903007816 to R.C.M.). R. C. M. received a Université de Paris COVID-19 grant. A. L. is also supported by ANR-17-CE14-0002-01, ANR-19-CE14-0020, Fondation Francophone pour la Recherche sur le Diabète. A. L., C. B., A. V.-P. and JF. G. are supported by RHU QUID-NASH (ANR-17-RHUS-009); M. R. and L.Ber. were supported by French Ministry of Research grants, A.T. and Z.G. were supported by RHU QUID-NASH; C. R. was supported by Fondation Francophone contre le Diabète, P. S. was supported by Juvenile Diabetes Research Foundation. Servier Medical Art for the free medical images (licensed under a Creative Commons Attribution 3.0 Unported License).

## Authors contribution

H. F. collected most of the samples, clinical data and performed experiments; M. R. performed experiments and all bioinformatics analyses. L. Bea. performed experiments, analyzed data and managed experimental procedures. L. Ber analyzed data. A. T. and S. Le. performed in vitro experiments. S. Lu. collected samples. Z. G., C. R. and P. S. performed flow cytometry experiments and analyses. M.H.N. performed CBA and K. B. and M. A. performed MSD Quickplex experiments. C. B., A. V.-P., JF. G., B. T., F. P., J. G., Y. Y. and JF. T. performed patient recruitment and analyzed clinical parameters. B.V. and D.D isolated and characterized SARS-CoV2. M. R., L. Bea., L. Ber., A. T., S. Le., H.F., R.C.M. and A. L. wrote the manuscript. H.F., L. Bea, R. C. M. and A. L. conceived the study. A. L. supervised the study. All authors edited and approved the manuscript.

## Conflict of interest statement

The authors declare no conflict of interest.

## Data availability statement

Data generated during this study will be available before publication in a repository. Access code will be given upon request.

## Code availability statement

Code used during this study will be available before publication in a repository. Access code will be given upon request.

## Notes

### Competing Interest Statement

The authors have declared no competing interest.

### Clinical Trial

This study did not aim to test any treatment.

### Author Declarations

The Ethics Committees approved clinical investigations. Informed consent was obtained from each enrolled patient. Patients from Bichat Hospital (Paris, France) were included in the French COVID cohort (NCT04262921). Ethics approval for this cohort was given on February 5th, 2020 by the French Ethics Committee CPP-Ile de France-VI (ID CRB: 2020-A00256-33). This cohort is sponsored by Inserm and supported by the REACTing consortium and by the French Ministry of Health (PHRC n20-0424). Samples from these patients were derived from samples collected in routine care. Patients from Cochin Hospital (Paris, France) were recruited in the setting of the local RADIPEM biological samples collection derived from samples collected in routine care. Biological collection and informed consent were approved by the Direction de la Recherche Clinique et Innovation (DRCI) and the French Ministry of Research (N2019-3677). Investigations with control patients from QUID-NASH were approved by Comite de Protection des Personnes de Sud Mediterranee (V) #18.021, N of QUID project registration: 2018-A00311-54.

## References

1. Wrapp, D. et al. Cryo-EM structure of the 2019-nCoV spike in the prefusion conformation. Science 367, 1260–1263 (2020).

2. Tay, M. Z., Poh, C. M., Rénia, L., MacAry, P. A. & Ng, L. F. P. The trinity of COVID-19: immunity, inflammation and intervention. Nature Reviews Immunology 1-12 (2020) doi:10.1038/s41577-020-0311-8.

3. Chen, N. et al. Epidemiological and clinical characteristics of 99 cases of 2019 novel coronavirus pneumonia in Wuhan, China: a descriptive study. Lancet 395, 507–513 (2020).

4. Huang, C. et al. Clinical features of patients infected with 2019 novel coronavirus in Wuhan, China. Lancet 395, 497–506 (2020).

5. Thompson, B. T., Chambers, R. C. & Liu, K. D. Acute Respiratory Distress Syndrome. N. Engl. J. Med. 377, 562–572 (2017).

6. Matthay, M. A. et al. Acute respiratory distress syndrome. Nature Reviews Disease Primers 5, 1–22 (2019).

7. Gorbalenya, A. E. et al. The species Severe acute respiratory syndrome-related coronavirus: classifying 2019-nCoV and naming it SARS-CoV-2. Nature Microbiology 5, 536–544 (2020).

8. Crosby, C. M. & Kronenberg, M. Tissue-specific functions of invariant natural killer T cells. Nat Rev Immunol 18, 559–574 (2018).

9. McCarthy, N. E. & Eberl, M. Human yô T-Cell Control of Mucosal Immunity and Inflammation. Front. Immunol. 9, (2018).

10. Toubal, A., Nel, I., Lotersztajn, S. & Lehuen, A. Mucosal-associated invariant T cells and disease. Nat. Rev. Immunol. 19, 643–657 (2019).

11. Trottein, F. & Paget, C. Natural Killer T Cells and Mucosal-Associated Invariant T Cells in Lung Infections. Front Immunol 9, (2018).

12. Constantinides, M. G. et al. MAIT cells are imprinted by the microbiota in early life and promote tissue repair. Science 366, (2019).

13. Leng, T. et al. TCR and Inflammatory Signals Tune Human MAIT Cells to Exert Specific Tissue Repair and Effector Functions. Cell Rep 28, 3077-3091.e5 (2019).

14. Stehle, C., Hernández, D. C. & Romagnani, C. Innate lymphoid cells in lung infection and immunity. Immunol. Rev. 286, 102–119 (2018).

15. Vivier, E. et al. Innate Lymphoid Cells: 10 Years On. Cell 174, 1054–1066 (2018).

16. Treiner, E. et al. Selection of evolutionarily conserved mucosal-associated invariant T cells by MR1. Nature 422, 164–169 (2003).

17. Corbett, A. J. et al. T-cell activation by transitory neo-antigens derived from distinct microbial pathways. Nature 509, 361–365 (2014).

18. Loh, L. et al. Human mucosal-associated invariant T cells contribute to antiviral influenza immunity via IL-18-dependent activation. Proc. Natl. Acad. Sci. U.S.A. 113, 10133–10138 (2016).

19. Ussher, J. E., Willberg, C. B. & Klenerman, P. MAIT cells and viruses. Immunology & Cell Biology 96, 630–641 (2018).

20. van Wilgenburg, B. et al. MAIT cells are activated during human viral infections. Nat Commun 7, 11653 (2016).

21. Ussher, J. E. et al. CD161++CD3+ T cells, including the MAIT cell subset, are specifically activated by IL-12+IL-18 in a TCR-independent manner. Eur J Immunol 44, 195–203 (2014).

22. Barathan, M. et al. Peripheral loss of CD8(+) CD161(++) TCRVα7-2(+) mucosal-associated invariant T cells in chronic hepatitis C virus-infected patients. Eur. J. Clin. Invest. 46, 170–180 (2016).

23. Leeansyah, E. et al. Activation, exhaustion, and persistent decline of the antimicrobial MR1-restricted MAIT-cell population in chronic HIV-1 infection. Blood 121, 1124–1135 (2013).

24. Leeansyah, E. et al. Arming of MAIT Cell Cytolytic Antimicrobial Activity Is Induced by IL-7 and Defective in HIV-1 Infection. PLoS Pathog. 11, e1005072 (2015).

25. Wang, F. et al. Characteristics of Peripheral Lymphocyte Subset Alteration in COVID-19 Pneumonia. J. Infect. Dis. 221, 1762–1769 (2020).

26. Silvin, A. et al. Elevated calprotectin and abnormal myeloid cell subsets discriminate severe from mild COVID-19. Cell (2020) doi:10.1016/j.cell.2020.08.002.

27. Schulte-Schrepping, J. et al. Severe COVID-19 is marked by a dysregulated myeloid cell compartment. Cell (2020) doi:10.1016/j.cell.2020.08.001.

28. Hadjadj, J. et al. Impaired type I interferon activity and inflammatory responses in severe COVID-19 patients. Science (2020) doi:10.1126/science.abc6027.

29. Zhang, J.-Y. et al. Single-cell landscape of immunological responses in patients with COVID-19. Nat. Immunol. (2020) doi:10.1038/s41590-020-0762-x.

30. Jouan, Y. et al. Functional alteration of innate T cells in critically ill Covid-19 patients. *medRxiv* 2020.05.03.20089300 (2020) doi:10.1101/2020.05.03.20089300.

31. Chua, R. L. et al. COVID-19 severity correlates with airway epithelium-immune cell interactions identified by single-cell analysis. Nat. Biotechnol. 38, 970–979 (2020).

32. Dusseaux, M. et al. Human MAIT cells are xenobiotic-resistant, tissue-targeted, CD161hi IL-17-secreting T cells. Blood 117, 1250–1259 (2011).

33. Hegde, P. et al. Mucosal-associated invariant T cells are a profibrogenic immune cell population in the liver. Nat Commun 9, 2146 (2018).

34. Rouxel, O. et al. Cytotoxic and regulatory roles of mucosal-associated invariant T cells in type 1 diabetes. Nat. Immunol. 18, 1321–1331 (2017).

35. Magalhaes, I. et al. Mucosal-associated invariant T cell alterations in obese and type 2 diabetic patients. J. Clin. Invest. 125, 1752–1762 (2015).

36. Toubal, A. et al. Mucosal-associated invariant T cells promote inflammation and intestinal dysbiosis leading to metabolic dysfunction during obesity. Nat Commun 11, 3755 (2020).

37. Zhou, F. et al. Clinical course and risk factors for mortality of adult inpatients with COVID-19 in Wuhan, China: a retrospective cohort study. Lancet 395, 1054–1062 (2020).

38. Cariou, B. et al. Phenotypic characteristics and prognosis of inpatients with COVID-19 and diabetes: the CORONADO study. Diabetologia 63, 1500–1515 (2020).

39. Lisco, G. et al. Hypothesized mechanisms explaining poor prognosis in type 2 diabetes patients with COVID-19: a review. Endocrine (2020) doi:10.1007/s12020-020-02444-9.

40. Czernichow, S. et al. Obesity doubles mortality in patients hospitalized for SARS-CoV-2 in Paris hospitals, France: a cohort study on 5795 patients. Obesity (Silver Spring) (2020) doi:10.1002/oby.23014.

41. Grimaldi, D. et al. Specific MAIT cell behaviour among innate-like T lymphocytes in critically ill patients with severe infections. Intensive Care Med 40, 192–201 (2014).

42. Cosgrove, C. et al. Early and nonreversible decrease of CD161++ /MAIT cells in HIV infection. Blood 121, 951–961 (2013).

43. Ishimori, A. et al. Circulating activated innate lymphoid cells and mucosal-associated invariant T cells are associated with airflow limitation in patients with asthma. Allergology International 66, 302–309 (2017).

44. Lei, X. et al. Activation and evasion of type I interferon responses by SARS-CoV-2. Nat Commun 11, 3810 (2020).

45. Dias, J., Leeansyah, E. & Sandberg, J. K. Multiple layers of heterogeneity and subset diversity in human MAIT cell responses to distinct microorganisms and to innate cytokines. Proc Natl Acad Sci U S A 114, E5434-E5443 (2017).

46. Leng, T. et al. TCR and Inflammatory Signals Tune Human MAIT Cells to Exert Specific Tissue Repair and Effector Functions. Cell Reports 28, 3077-3091.e5 (2019).

47. Blackley, S. et al. Primary human splenic macrophages, but not T or B cells, are the principal target cells for dengue virus infection in vitro. J. Virol. 81, 13325–13334 (2007).

48. Hasnain, S. E. et al. Host-pathogen interactions during apoptosis. J. Biosci. 28, 349–358 (2003).

49. Gordon, D. E. et al. A SARS-CoV-2 protein interaction map reveals targets for drug repurposing. Nature 583, 459–468 (2020).

50. Symons, J. A., Alcamí, A. & Smith, G. L. Vaccinia virus encodes a soluble type I interferon receptor of novel structure and broad species specificity. Cell 81, 551–560 (1995).

51. Désiré, N. et al. Quantification of human immunodeficiency virus type 1 proviral load by a TaqMan real-time PCR assay. J. Clin. Microbiol. 39, 1303–1310 (2001).

52. Nowicka, M. et al. CyTOF workflow: differential discovery in high-throughput high-dimensional cytometry datasets. F1000Res 6, 748 (2017).

